# The association between outdoor ambient temperature and depression and mania: an ecological momentary assessment study

**DOI:** 10.1101/2024.10.10.24315229

**Authors:** P Clery, JF Hayes, N Launders, R Thompson, A Kandola, DPJ Osborn, EL Lawrance, A Jeffery, J Dykxhoorn

**Affiliations:** Division of Psychiatry, University College London, London, UK; North London Mental Health Partnership, London, UK; Juli Health, Hull, Massachusetts, USA; Department of Epidemiology and Biostatistics, School of Public Health, Imperial College London, London, UK; NIHR School for Public Health Research, England, UK; MRC Unit of Lifelong Health and Ageing, UCL, UK; Climate Cares Centre, Institute of Global Health Innovation, Imperial College London, London, UK; Grantham Institute for Climate Change and the Environment, Imperial College London, London, UK

## Abstract

**Background:** Environmental heat exposure can negatively impact mental health. Evidence for its effect on mood disorder symptoms is inconsistent. Current studies are limited by poor temporal and geographical resolution.

**Methods:** We used ecological momentary assessment (EMA) data from the smartphone app *juli* to investigate the association between real-time mean and maximum ambient temperature collected from smartphone geolocation, and depressive and manic symptom scales, every two weeks, in adults with depression and bipolar disorder. We used negative binomial mixed-effects regression models, controlled for demographic and weather variables, and stratified by season.

**Results:** We analysed data from 4,000 participants with depressive symptom scores and 2,132 with manic symptom scores, between 2021 and 2023. We found that each 1°C increase in mean daily temperature in the preceding two weeks was associated with a 0.2% reduction in depressive symptom scores (coeff 0.998, 95%CI 0.997-0.999) and a 0.4% increase in manic symptom scores (coeff 1.004, 95%CI 1.001-1.007). Associations between maximum temperature and symptom scores followed a similar pattern.

**Limitations:** We were unable to capture several socio-demographic covariates, had limited geographical information due to privacy regulations, and included a non-random sample.

**Conclusions:** We found evidence that higher temperatures were associated with increased manic symptoms and decreased depressive symptoms, indicating an important relationship between temperature and the mood disorder continuum. With global heating, there is a need to understand the impact of temperature on mood symptoms, to provide targeted clinical prevention and support. This study demonstrates potential for EMA methods to inform our understanding of these links.

## Introduction

Evidence suggests environmental heat exposure negatively impacts mental health (Lawrance et al., 2022; Lee et al., 2023). Recent systematic reviews show higher ambient temperatures are associated with worse mental health outcomes, from increased rates of suicide and hospital admission to poorer mental wellbeing (Thompson et al., 2018; Gao et al., 2019; Heo et al., 2021; Liu et al., 2021; Thompson & Lawrance et al., 2023). The differential burden across mental health conditions is not clear. In the mood disorder population, most evidence explores hospitalisation or mortality outcomes. There is a paucity of evidence for the effect of temperature on less severe clinical outcomes in a clinical population of people with mood disorders. With climate change projected to increase global temperatures (IPCC, 2023), and people with pre-existing mental health conditions experiencing a greater risk of health impacts during hotter temperatures (Woodland et al., 2023), it is important to consider the relationship between temperature and depressive and manic symptoms.

Several potential overlapping biological, cognitive, and socioeconomic pathways between temperature and mood have been proposed (Berry et al., 2010; Lõhmus, 2018). Some studies explore *extreme* heat, whilst others explore a temperature continuum. Evidence shows an increase in violence and agitation at higher temperatures (Tiihonen et al., 2017), suggesting heat may impact affect and agitation. The serotonin theory linking heat and mood could contribute to both elevated and depressed mood across both mild and extreme heat (Lambert et al., 2002; Lowry et al., 2009). In extreme heat, physiological and cognitive effects of heat stress (Kovats and Hajat, 2008), sleep disturbance (Mullins and White, 2019), and impaired thermoregulation due to psychotropic medication (Martin-Latry et al., 2007; Stöllberger et al., 2009) may all cause an exacerbation of mood disorder symptoms. Further, reduced socioeconomic capacity for adaptation or mitigation against extreme heat (Deschênes and Greenstone, 2011; Zivin and Neidell, 2014) and the impact of extreme heat on the economy (Oppermann et al., 2021) can impact on mood disorder symptoms indirectly via social determinants of mental health.

Global findings on the relationship between temperature and depressive and manic symptoms are inconsistent. Some studies suggest hotter average temperatures may increase the incidence and symptom severity of depression (Chen et al., 2019; Henríquez-Sánchez et al., 2014; Hou et al., 2023; Wang et al., 2023, 2014; Zhou et al., 2023) and mania (Bullock et al., 2017; Geoffroy et al., 2014; Medici et al., 2016; Montes et al., 2021; Shapira et al., 2004; Sung et al., 2013). Some report no consistent effect (McWilliams et al., 2014; Pailler and Tsaneva, 2018; Xue et al., 2019). An increase in negative emotions, stress and fatigue are reported at temperatures above 32°C (Noelke et al., 2016), whilst a depressive effect on general mood and wellbeing has been demonstrated in colder climates (Davis and Lowell, 2002; Kessler et al., 2007). Factors such as sunlight hours and outdoor activity (Kim et al., 2019), gender, age, and socioeconomic ability to mitigate against effects of extreme heat (Bouchama et al., 2007; Thompson & Lawrance et al., 2023) could account for differences.

Discrepancies also arise due to methodological limitations. The current literature comprises heterogeneity in outcome measures, misclassification bias of temperature exposure, time-lag effects, and lack of adjustment for weather confounders (Hwong et al., 2022; Massazza et al., 2022; Thompson & Lawrance et al., 2023). Studies use large population datasets to detect small changes, whereby attaining robust and meaningful environmental and clinical variables in a relevant population can be difficult. Of the recent studies measuring the effect of temperature on depressive symptoms, none studied a mood disorder clinical population, four used the same two Chinese datasets (Hou et al., 2023; Jin et al., 2023; Wang et al., 2023; Xue et al., 2019) and three were conducted on adults aged over 65 only (Jin et al., 2023; O’Hare et al., 2016; Wang et al., 2023).

To overcome some of these methodological limitations, a more ideal approach would use temperature measures representative of an individual’s exposure, validated clinical symptom scales, and repeated data collection with short time-lags between exposure and outcome. Ecological momentary assessment (EMA) offers an approach to address some of these. EMA repeatedly samples participant behaviour, activity, and psychological processes in real-time in their naturalistic environment (Shiffman et al., 2008). It has been used to investigate the effect of environmental variables such as air pollution (Kandola and Hayes, 2023) and nature-based surroundings (Bakolis et al., 2018; Bergou et al., 2022) on mental health outcomes. To our knowledge, only one study to date has used EMA to investigate the association between temperature and mood on a small sample in Switzerland (Bundo et al., 2023).

Our study aimed to use EMA data from the digital health platform and smartphone application *juli* to investigate the association between short-term real-time ambient outdoor temperature, and two-weekly depressive and manic symptoms in a large clinical cohort of adults with depression or bipolar affective disorder. Our study investigates depressive and manic symptom scales separately. This is for clinical applicability, as the same clinical criteria are used to diagnose depressive episodes in both unipolar and bipolar depression (World Health Organization, 2022), and because we hypothesise the effects of temperature to vary by symptoms rather than by disorder. We hypothesised higher mean and maximum temperatures would be associated with increased depressive and manic symptom scores, and there would be a stronger association in males and older age groups.

## Methods

### Study design and participants

We used data extracted from *juli* (juli Health, 2022), a smartphone app for iOS and Android designed to support patients with chronic conditions to track symptoms and environmental conditions which affect their health. Participants were included in the study if they reported physician-diagnosed depression or bipolar disorder and if they were active users of *juli* between July 2021 and March 2023. All participants were over 18 years old and consented to their aggregated de-identified data being used for research. Ethical approval for the study was provided by the University College London Research Ethics Committee (ID 19413/002). We pre-registered the protocol on Open Science Framework (Clery et al., 2023).

### Outcomes

This study explored depressive symptoms and manic symptoms. Depressive symptoms were assessed using the Patient Health Questionnaire-8 (PHQ-8; Kroenke et al., 2009). The PHQ-8 is a widely-used clinical screening and research tool (Farber et al., 2020) and has been successfully used in EMA smartphone studies (Colombo et al., 2019). The more widely used PHQ-9 questionnaire (Moriarty et al., 2015) includes a suicidality question that is not included in the PHQ-8 due to the ethical and safety concerns with a positive response when collecting data digitally. The PHQ-8 asks participants eight questions about symptoms of depression over the previous two weeks. Scores range from 0 to 24; greater than 10 suggests major depressive disorder. App users with a diagnosis of depression or bipolar disorder were invited to complete the PHQ-8 every two weeks. Participants who had completed at least one PHQ-8 were included in the depressive symptoms cohort.

Manic symptoms were measured using the Altman Self Rating Mania scores (ASRM; Altman et al., 1997). The ASRM is a five-item, five-point self-reported scale with high clinical utility (Cerimele et al., 2019) and has been previously used in EMA smartphone studies (Tatham et al., 2022). The scale includes questions about elevated mood, self-confidence, sleep patterns, speech and activity level over the previous week. Scores range from 0 to 20; a score higher than five indicates mania (Altman et al., 1997). Participants with a diagnosis of bipolar disorder were invited to complete the ASRM every two weeks. Participants who had completed at least one ASRM were included in the manic symptoms cohort.

### Exposures

Mean and maximum outdoor ambient air temperature in degrees Celsius (°C) were measured during the two weeks prior to completing the PHQ-8 or ASRM. This time period was chosen because the symptom questionnaires ask “Over the last 2 weeks, how often were you bothered by…”. A mean temperature of all recordings over the two weeks was calculated. Temperature was estimated using data provided to *juli* from an Application Programming Interface software called *ambee* (2017). This software collects meteorological weather data for participant-specific smartphone geolocation on a 5-by-5km geospatial grid resolution globally. Participant smartphone geolocation was used to passively collect environmental data through *ambee*, then geolocation data was disposed of to ensure de-identification due to privacy regulations.

### Covariates

Gender (female, male, and trans-gender or non-binary), humidity (mean of daily humidity (%)), sunshine (mean counts of when weather was recorded as ‘clear’(n)) and air pollution (mean air quality index (AQI)) were included as confounders in our adjusted analysis. Measures of humidity, sunshine, and air pollution were calculated based on mean values in the two weeks prior to completing the PHQ-8 or ASRM. Gender (categories as above) and age (<45; ≥45 years old; excluding missing data) were included in interaction tests.

Covariates in sensitivity analysis included: an individual’s temperature norm, age (continuous variable) and step count. Temperature norms were defined as an individual’s annual and seasonal mean temperature (in degrees Celsius (°C)) during the study period. We estimated the annual mean temperature as a mean of all the temperature measures provided by the participant in the dataset. We estimated the seasonal mean temperature as a mean of all the temperature measures provided by the participant in the dataset per each season. Step count was an ordinal measure (<1,000 steps; 1,000-3,000; 3,001-5,000; 5,001-7000; 7,001-9,000; >9,000) with groupings based on real-world applicability of step count and to ensure sufficient sample size in each group. We adjusted for step count as a proxy for an individual’s activity and thus potential exposure to outdoor ambient temperature.

### Statistical analysis

#### Primary analysis

PHQ-8 nor ASRM scores did not meet linear assumptions required for linear regression (supplement A). Negative binomial regression models were found to be a better fit (supplement A). Although negative binomial models are typically used for count variables, our symptom score data have the same characteristics of discreet points, and we will interpret the outcome as a percent change in symptom scores. As participants were able to complete symptom scales multiple times during the study period, we used mixed effects models with random effects to account for the clustering within individuals.

We estimated unadjusted and adjusted estimates for the overall association between mean and maximum temperature and depressive symptoms and manic symptoms. We adjusted for gender, humidity, sunshine, and air pollution. Then, we stratified the analysis by season (spring, summer, autumn, and winter), generating unadjusted and adjusted estimates across each stratum. We calculated the exponentiated coefficient and generated 95% confidence intervals (CI) from negative binomial models, representing the percent change in symptom scores associated for each 1⁰C increase in temperature. All analyses were conducted in R Version 4.2.1 (R Core Team, 2022) using the glmmTMB package (Brooks et al., 2017).

#### Missing data and sensitivity analyses

We were unable to impute missing data as there were insufficient complete variables for imputation. Therefore, analysis was conducted on complete cases, excluding participants missing information for outcomes, exposures, or covariates. We compared participants with complete case data versus missing data using logistic regression and report the odds ratio (OR) with 95% confidence intervals.

We fitted interaction terms on the adjusted model because we hypothesised that the association may differ by gender and age. We performed several sensitivity analyses with covariates that contained too much missing data to be included in main adjusted models. We included separately individual temperature norm, age, and step count to the adjusted models. We conducted additional analysis of the results by diagnosis (depression vs. bipolar disorder) by fitting an interaction term in the adjusted model. This additional analysis was not specified in our pre-registered protocol (Clery et al., 2023), but was included following further discussions with co-authors.

## Results

### Descriptive statistics

#### Participants

The total number of participants with physician-diagnosed depression or bipolar disorder was 9,771. The number of participants who completed at least one PHQ-8 was 7,268. Of these, 70% (n=5,107) provided temperature readings in the two weeks preceding the PHQ-8. This comprised the depressive symptoms cohort. Of these, 55% (n=4,000) had complete data on confounder covariates (gender, humidity and air quality). This comprised the depressive symptoms cohort with complete cases.

The number of participants who completed at least one ASRM was 3,331. Of these, 75% (n=2,686) provided temperature readings within two weeks preceding the ASRM. This comprised the manic symptoms cohort. Of these, 79% (n=2,132) had complete data on confounder covariates. This comprised the manic symptoms cohort with complete cases (Figure 1).

**Figure 1.**
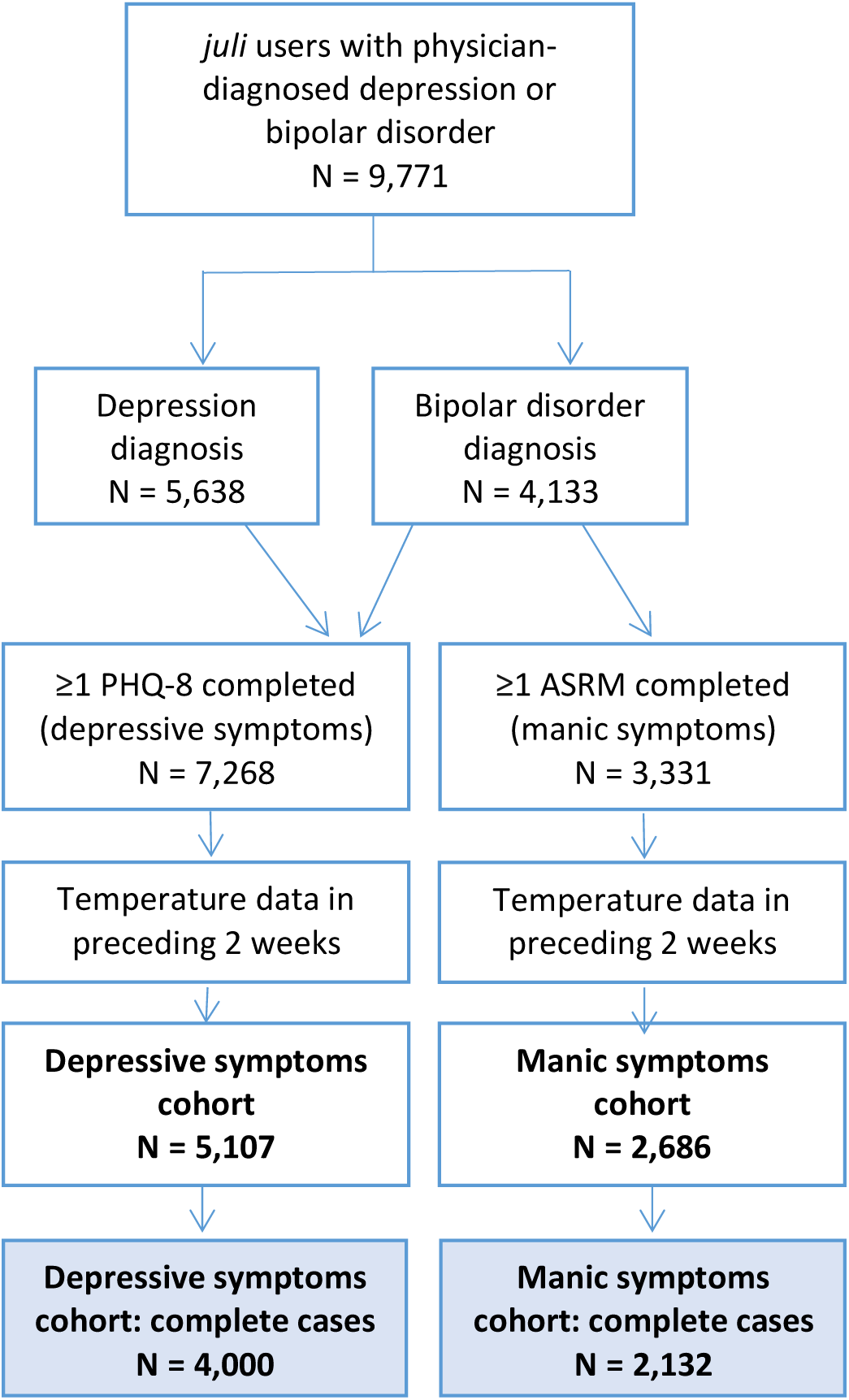
Participant flowchart.

#### Baseline characteristics

In both the depressive symptoms and manic symptoms cohorts, most participants were diagnosed more than five years before the study start, were female, were White ethnicity and had a mean age of 30.9 (SD 11.8) and 30.4 (SD 10.5), respectively (Table 1). Most participants were from North America (84%; supplement B). There were 1,107 (22%) participants in the depressive symptoms cohort and 554 (21%) participants in the manic symptoms cohort with missing data on at least one covariate. Participants with missing data were younger, were less likely to have physical health comorbidities, and had a shorter duration of their mental health diagnosis (supplement C).

**Table 1:** Baseline Characteristics (complete case*)

| Characteristic | Depressive Symptoms Cohort |  | Manic Symptoms Cohort |  |
| --- | --- | --- | --- | --- |
|  | *n* | % | *n* | % |
| **Total** | 4,000 | 100.0 | 2,132 | 100.0 |
| **Gender\*** |  |  |  |  |
| Female | 3,178 | 79.4 | 1,710 | 80.2 |
| Male | 539 | 13.5 | 289 | 13.6 |
| Non-binary or Transgender | 283 | 7.1 | 133 | 6.2 |
| **Age (years)** |  |  |  |  |
| Mean (SD) | Mean: 30.93 | SD: 11.81 | Mean: 30.45 | SD: 10.45 |
| **Ethnicity** |  |  |  |  |
| American Indian, Alaska Native, Native Hawaiian, or other Pacific Islander | 16 | 0.4 | 16 | 0.8 |
| Asian | 40 | 1.0 | 17 | 0.8 |
| Black or African American | 110 | 2.8 | 68 | 3.2 |
| White | 942 | 23.5 | 538 | 25.2 |
| Not stated | 2,892 | 72.3 | 1,493 | 70.0 |
| **Hispanic** |  |  |  |  |
| No | 1,016 | 25.4 | 586 | 27.5 |
| Yes | 137 | 3.4 | 89 | 4.2 |
| Not stated | 2,847 | 71.2 | 1,457 | 68.3 |
| **Diagnosis** |  |  |  |  |
| Depression | 2,242 | 56.0 | 0 | 0.0 |
| Bipolar Disorder | 1,758 | 44.0 | 2,132 | 100.0 |
| **Duration of Diagnosis** |  |  |  |  |
| < 1 year | 356 | 8.9 | 260 | 12.2 |
| 1-5 years | 1,261 | 31.5 | 703 | 33.0 |
| > 5 years | 2,383 | 59.6 | 1,169 | 54.8 |
| **Physical health comorbidity<sup>‡</sup>** | 602 | 15.0 | 157 | 7.4 |
| **Temperature (daily mean °C)\*** |  |  |  |  |
| Mean (SD) | Mean: 14.22 | SD: 8.12 | Mean: 15.50 | SD: 7.95 |
| **Humidity (daily mean %)\*** |  |  |  |  |
| Mean (SD) | Mean: 65.78 | SD: 13.41 | Mean: 63.86 | SD: 13.68 |
| **Air quality (daily mean AQI)\*** |  |  |  |  |
| Mean (SD) | Mean: 45.84 | SD: 22.42 | Mean: 41.81 | SD: 16.06 |
| **Sunshine (daily mean count)\*** |  |  |  |  |
| Mean (SD) | Mean: 6.08 | SD: 5.29 | Mean: 6.23 | SD: 5.98 |
| **Step count (daily mean steps)** |  |  |  |  |
| <1,000 | 350 | 8.8 | 173 | 8.1 |
| 1,000-3,000 | 1,358 | 34.0 | 750 | 35.2 |
| 3,001-5,000 | 1,046 | 26.2 | 527 | 24.7 |
| 5,001-7,000 | 523 | 13.1 | 274 | 12.9 |
| 7,001-9,000 | 278 | 7.0 | 144 | 6.8 |
| >9000 | 163 | 4.1 | 97 | 4.5 |
| Not recorded | 282 | 7.0 | 167 | 7.8 |

\* The star marks the variables used for complete cases - participants included in the complete case analysis had information on gender, temperature, humidity, air quality
<sup>‡</sup> Comorbidity of: asthma, migraines, non-migraine headaches, chronic pain or hypertension

#### Depressive and manic symptom scores

Participants completed between one and 37 PHQ-8 and ASRM questionnaires. In the depressive symptoms cohort, the mean number of PHQ-8s completed was 2.61 (SD 3.44). In total 10,423 PHQ-8 questionnaires were completed. The mean PHQ-8 score was 12.18 (SD 6.23), meeting the threshold for clinical depressive disorder (cut-off score is 10).

In the manic symptoms cohort, the mean number of ASRMs completed was 2.44 (SD 3.36). In total, 5,204 ASRM questionnaires were completed. The mean ASRM score was 4.29 (SD 3.90), indicating high levels of symptoms but not meeting criteria for mania (cut-off score is 5).

Completion was highest in the winter season in the depressive symptoms cohort, and highest in summer in the manic symptoms cohort. Completion was lowest in spring for both cohorts (supplement D).

#### Temperature

In the depressive symptoms cohort, participants contributed between one and 616 temperature recordings. The mean number of recordings was 28.31 (SD 50.45) and in total 113,245 recordings were contributed. The mean temperature was 14.22⁰C (SD 8.12). The lowest recorded temperature was −35⁰C and highest recorded temperature was 46⁰C.

In the manic symptoms cohort, participants contributed between one and 614 temperature recordings. The mean number of recordings was 25.03 (SD 49.15) and in total 53,366 recordings were contributed. The mean temperature was 15.50⁰C (SD 7.95). The lowest recorded temperature was −34⁰C and highest recorded temperature was 45⁰C (supplement D).

### Association between mean and maximum temperature and depressive symptoms

#### Association between 14-day mean daily temperature and depressive symptoms

We found that each 1°C increase in mean temperature was associated with a 0.2% reduction in depressive symptoms (coeff 0.998, 95%CI 0.997-0.999). Unadjusted and fully adjusted effect estimates were the same (Table 2).

**Table 2:**
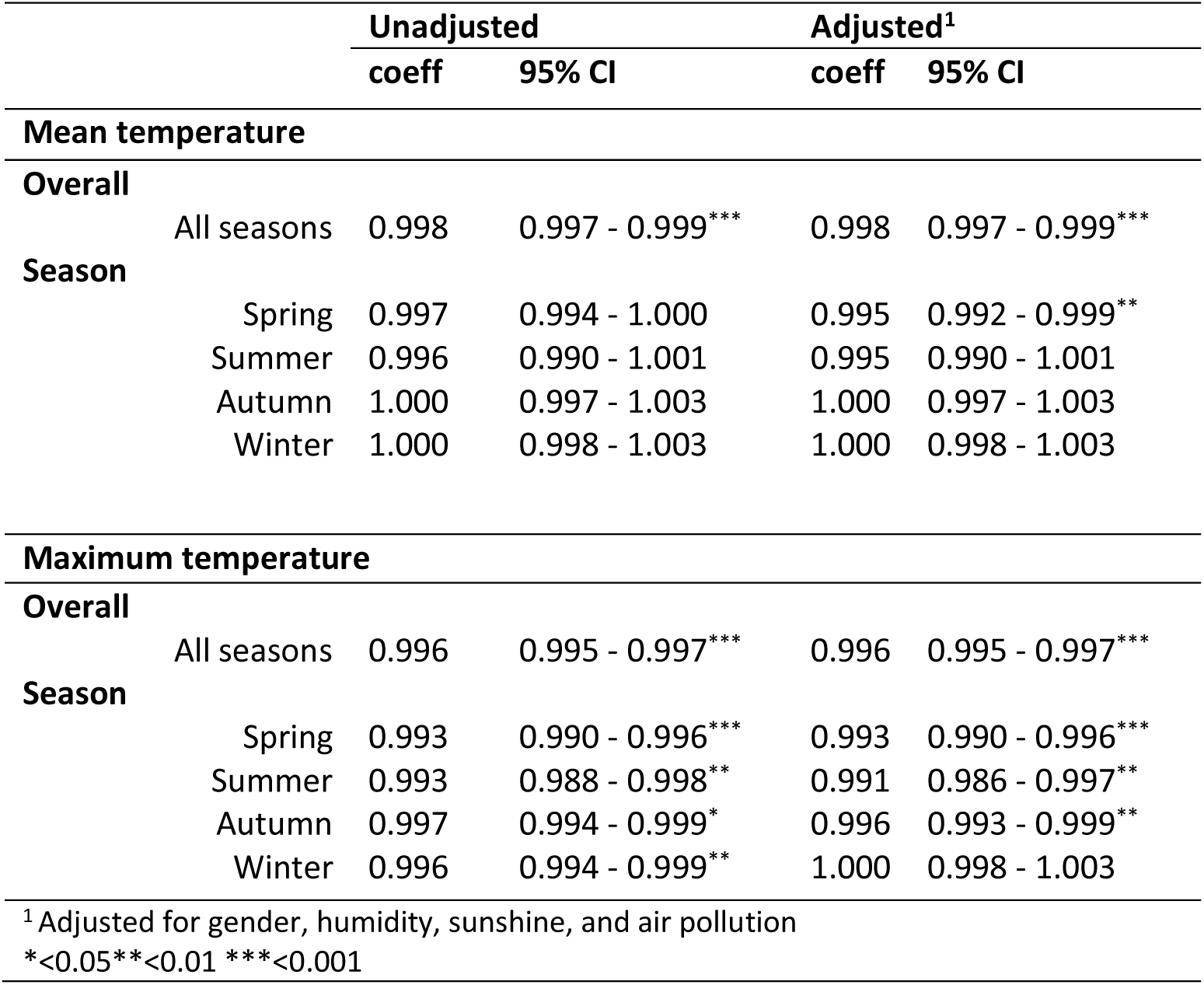
Association between temperature (mean, maximum) and depressive symptoms (PHQ-8)

When stratified by season, the association was most pronounced in spring (percent change: 0.5%; coeff: 0.995, 95%CI 0.992-0.999), with a similar trend observed for summer, although the association crossed the null (percent change: 0.5%; coeff 0.995, 95%CI 0.990-1.001). We found no evidence of an association between mean daily temperature and depressive symptoms in autumn or winter (percent change: 0.0%; coeff 1.000, 95%CI 0.997-1.003; coeff 1.000, 95%CI 0.998-1.003, respectively; Table 2).

#### Association between 14-day maximum temperature and depressive symptoms

We observed an association between increased maximum temperature and lower depressive symptoms (percent change: 0.4%; coeff 0.996, 95%CI 0.995-0.997). This pattern was observed for all seasons, except winter (Table 2).

### Association between mean and maximum temperature and manic symptoms

#### Association between 14-day mean daily temperature and manic symptoms

We found that, each 1°C increase in daily mean temperature across all seasons was associated with a 0.4% increase in manic symptom scores (coeff 1.004, 95%CI 1.001-1.007). When stratified by season, the association was largest and strongest in autumn, where each 1°C increase corresponded to a 1.1% increase in manic symptom scores (coeff 1.011, 95%CI 1.002-1.021) (Table 3).

**Table 3:** Association between temperature (mean, maximum) and manic symptoms (ASRM)

|  |  | Unadjusted |  | Adjusted<sup>1</sup> |  |
| --- | --- | --- | --- | --- | --- |
|  |  | coeff | 95% CI | coeff | 95% CI |
| **Mean temperature** |  |  |  |  |  |
| **Overall** |  |  |  |  |  |
|  | All seasons | 1.004 | 1.001 - 1.007** | 1.004 | 1.001 - 1.007** |
| **Season** |  |  |  |  |  |
|  | Spring | 1.005 | 0.997 - 1.013 | 1.002 | 0.994 - 1.010 |
|  | Summer | 0.999 | 0.987 - 1.012 | 1.000 | 0.986 - 1.014 |
|  | Autumn | 1.011 | 1.002 - 1.020* | 1.011 | 1.002 - 1.021* |
|  | Winter | 1.004 | 0.996 - 1.012 | 1.004 | 0.996 - 1.012 |
| **Maximum temperature** |  |  |  |  |  |
| **Overall** |  |  |  |  |  |
|  | All seasons | 1.004 | 1.001 - 1.006* | 1.003 | 1.000 - 1.007* |
| **Season** |  |  |  |  |  |
|  | Spring | 1.006 | 0.998 - 1.013 | 1.003 | 0.995 - 1.011 |
|  | Summer | 0.997 | 0.985 - 1.009 | 0.997 | 0.983 - 1.011 |
|  | Autumn | 1.006 | 0.998 - 1.014 | 1.007 | 0.999 - 1.016 |
|  | Winter | 1.001 | 0.994 - 1.009 | 1.002 | 0.994 - 1.010 |
| <sup>1</sup> Adjusted for gender, humidity, sunshine, and air pollution |  |  |  |  |  |
| * < 0.05 ** < 0.01 *** < 0.001 |  |  |  |  |  |

#### Association between 14-day maximum temperature and manic symptoms

We found that a 1°C increase in maximum temperature across all seasons corresponded to a 0.3% increase in manic symptom scores (coeff 1.003, 95%CI 1.000 to 1.007). When stratified by season, trends were similar across all seasons except summer, but all associations crossed the null (Table 3).

### Sensitivity analyses

#### Interaction effects

We did not detect an interaction for the effect of gender, age, or diagnosis, so we did not disaggregate results by these characteristics (supplement E).

#### Sensitivity analyses

We conducted sensitivity analysis where we further adjusted for individual temperature norms, age, and step count. We excluded participants missing age or step count, therefore, n= 2,486 and n=3,710 respectively were included in the depressive cohort for this sensitivity analysis and n=1,389 and n=1,958 respectively were included in the sensitivity analysis of the manic cohort (supplement E).

#### Depressive symptoms

The overall association between *mean* daily temperature and depressive symptoms scores appeared similar following additional adjustment for temperature norms (percent change: 0.2%; coeff 0.998, 95%CI 0.997-1.000) but we found some differences by season. We found that depression scores in spring and summer decreased by 0.9% for each 1°C increase (spring coeff 0.991, 95%CI 0.986-0.996; summer coeff 0.991, 95%CI 0.983-0.999). In winter, we found the opposite association compared with primary analyses, where each 1°C increase in mean temperature was associated with a 0.5% increase in depressive symptoms (coeff 1.005, 95%CI 1.000-1.009). When we adjusted the association for *maximum* temperature by individual temperature norms, we found stronger evidence of a reduction in depressive symptoms with increased temperature. We found a 1.1% reduction in depressive symptoms for each 1°C increase in the spring (coeff 0.989, 95%CI 0.985-0.993) and summer (coeff 0.989, 95%CI 0.983-0.995). These results also persisted after separate adjustment for age and step count (supplement E).

#### Manic symptoms

Adjusting for individual temperature norms in the manic symptoms cohort found similar results to the primary adjusted analysis, where each 1°C increase in *mean* daily temperature corresponded to a 0.7% increase in manic symptom scores (coeff 1.007, 95%CI 1.002-1.011) and each 1°C increase in *maximum* daily temperature corresponded to a 0.5% increase in manic symptom scores (coeff 1.005, 95%CI 1.001-1.009). When we stratified by season, there were was no evidence for an association. There was no evidence for an association when adjusting for age or step count (supplement E).

## Discussion

### Main findings

This novel EMA study found evidence that higher mean and maximum temperatures were associated with fewer depressive symptoms and more manic symptoms in people with a prior diagnosis of depression and bipolar disorder. Effect sizes were small such that for each 1°C increase, depressive symptom scores reduced by up to 1.1% and manic symptom scores increased by up to 1.9%. We found no differential effects across ages, genders or disorder.

### Comparison to previous literature

Our finding of reduced depressive symptoms with increased temperatures ran contrary to our *a priori* hypotheses. Prior studies show that higher temperatures are associated with worsening depressive symptoms and hospital admissions for mood disorders (Bundo et al., 2021; Burke et al., 2018; Chan et al., 2018; Hansen et al., 2008; Hou et al., 2023; Jin et al., 2023; Keller et al., 2005; Mullins and White, 2019; Trang et al., 2016; Wang et al., 2014; Yang et al., 2021; Zhang et al., 2020). However, these studies mostly looked at exposure to extreme heat or departure from ‘optimum’ temperature, thereby looking at extreme temperatures. The current study did not measure extreme heat, rather it studied the wide range of annual temperatures. This may explain why we did not find a positive relationship between temperature and depressive symptoms.

In contrast, some studies concur with our findings on depressed mood. These studies look at population measures of general mood and wellbeing. They show an improvement in general mood in warmer temperatures (Denissen et al., 2008; Jiang et al., 2022; Keller et al., 2005; Knapp et al., 2016). These results are therefore more in keeping with our study. However, these studies were not based on a clinical population and do not measure symptoms using clinical scales. They also do not focus on extreme heat. Taken together, the evidence suggests that higher temperatures within a mild temperate range may reduce depressive symptoms, whilst extreme high temperatures increase depressive symptoms. Alternatively, it may be possible that the current study is capturing a subclinical improvement in mood that is seen more generally on a population level, rather than a clinical change. The contradictory findings between hospital admission studies and population-based studies could also indicate an alternative explanation, that there is a different response to heat in people who are more severely and/or acutely unwell, compared with the general population and/or those with an established mood disorder diagnosis who are not acutely unwell. Indeed, a recent EMA study from Switzerland found that rising temperatures improved mood in the general population, but increased morbidity in those with a psychiatric disorder (Bundo et al., 2023).

Our findings support our *a priori* hypothesis that higher temperatures were associated with increased symptoms of mania i.e., elevated mood. This is consistent with large studies using seasonal pattern questionnaires and hospital admission data, which have found higher rates of hospital admissions for mania during hotter summer months (Geoffroy et al., 2014; Medici et al., 2016; Montes et al., 2021; Myers and Davies, 1978; Parker et al., 2017). Our results also echo findings from a smaller EMA study which found daily maximum temperatures predicted clinically-relevant mood changes, transitioning to mania (Bullock et al., 2017).

An interesting finding was that adjusting for an individual’s normal temperature exposure influenced the association between temperature and symptoms. This could indicate that the normal temperature that an individual is accustomed to may influence the association between the temperature and their mood disorder symptoms. Further, we found that this adjustment for individual temperature norms had a greater effect on the relationship between temperature and depressive symptoms, compared with the effect on manic symptoms. This could suggest higher temperatures have an impact on manic symptoms regardless of context, compared with depressive symptoms where contextual temperature norms matter more. Indeed, prior literature on depressive symptoms shows that deviance from optimum or normal temperature exposure for that individual (Jin et al., 2023), temperature variability (Wang et al., 2023) or a change in temperature (Xue et al., 2019), including rate or speed of change, may be more important than the temperature itself.

### Limitations

Several limitations should be noted, including restrictions to geographical and exposure information, selection bias of participants and incomplete measurement of key covariates.

Although we use precise geolocation, participant’s heat exposure may still not be accurate due to lack of information about artificial heating and cooling systems. Further, geolocation data were removed due to privacy regulations. Thus, we were unable to use geographic information to estimate temperature norms. For participants with few temperature estimates, or using the app more in a particular season, the calculated temperature norm may be inaccurate.

Differential patterns of app use based on symptoms or weather may also have affected our results. More PHQ-8s were completed during winter when depressive symptom scores were highest, suggesting participants may have used the app more frequently either when feeling low, or in the colder darker winter months, or both. There is currently no literature that has studied long-term seasonality trends of mental health-app usage (Dunster et al., 2021; Wu et al., 2021). Therefore, we are unable to compare our results to another sample. Trends of smartphone use in the United States suggest general mobile phone use shows no major seasonal shifts (Waber, 2014). We attempted to mitigate against the impact of seasonal effects on our results by stratifying by season and adjusting for temperature norms.

Participants may not be representative of the wider population of those diagnosed with depression and bipolar disorder. Participants were more likely to be well enough to participate, younger, female, have a smartphone, be interested in downloading the *juli* app and motivated to use it more than once. We are unable to comment on those who cannot use the app or do not have access to a smartphone. This may be related to certain relevant socioeconomic or disorder factors in this population, for example housing insecurity or compulsory psychiatric care.

Incomplete and inaccurate measurement of the confounders due to the nature of data collection may have also biased results. For example, when downloading the app, participants are asked to input their age using a scrolling menu, but a response was not mandatory. When we extracted the data, there was considerable missing data for age and unexpected patterns suggesting age may not be accurately reported (e.g., 10% of participants with age data were aged 18). Incomplete and potentially inaccurate data limit the interpretability of results of interaction effect by age group. This is an important consideration for studies using app-based data for research, as the way that data is requested by users can influence the accuracy of response.

Lastly, race and ethnicity variables were only added to baseline data collection half-way through study recruitment, whilst *juli* does not collect information on income, employment, education, urbanicity, or area-level deprivation. Other confounders of interest were missing for most participants (e.g., prescription medication and adherence, physical comorbidities, sleep, and hours spent outside) so we were not able to use them in analysis. Missingness across these variables prevented us from imputing because there was insufficient data to create plausible imputed datasets (Sterne et al., 2009). We therefore conducted complete case analyses, which reduced our sample size and may have biased our findings.

### Strengths

This study is the largest EMA study to date exploring the impact of climate and mental health in a clinical population, including over 4,000 people with depressive symptom data and 2,132 people with manic symptom data. Most notably, the EMA study design allowed us to overcome the methodological limitations of previous population studies which rely on exposures measured at a large spatial scale or at an ecological level (e.g., temperature data based on a participants registered post code or city). Our study was able to use an individual’s precise location and time-specific exposures, passively collected through their smartphone location. This also allowed us to adjust for humidity, sunshine and air pollution, which have been identified as important potential confounders in the relationship between temperature and mood disorders (Ding et al., 2016).

Further, we used standardised, validated and clinically relevant PHQ-8 and ASRM questionnaires to collect repeated outcome measurements of depressive and manic symptoms over two years. This detailed insight into depressive and manic symptoms allowed us to explore changes in symptom levels that may be undetectable when using hospital admission data or electronic health records, which are not likely to capture changes in mental health symptoms and are influenced by access to health care services and barriers to care.

### Clinical impact

Although our results indicate a small percent change in symptoms for each degree Celsius increase, small effects are common in this field and are of significant public health importance when considered at population level and over several degrees of temperature changes (e.g., during extreme heatwave events (Lenton et al., 2023)).

This study suggests it is important for mental health professionals and public mental health bodies to consider the impact of extreme heat events in people with bipolar disorder, as they are at risk of increased manic symptoms during this time. Importantly, this study has implications for mental health relapse prevention as it explores symptom changes in people who have not yet come to the attention of mental health services via hospital admission or mental health crisis. Understanding the effect of temperature on symptoms *before* reaching more severe outcomes of hospitalisation or mortality is invaluable for mental health services preparedness and targeted mental health support and interventions.

### Future directions

First, the different associations observed between temperature and depressive and manic symptoms suggests there may be different underlying mechanisms, and future research should explore the two symptom scales separately. Second, the apparent different relationships between symptoms and mild and extreme heat warrants exploring outcomes across the temperature range, including extreme heat only. Third, different temperature exposure measures may have non-linear relationships with mental health (Armstrong, 2006; Gasparrini et al., 2015). Although our data did not lend itself to a time series design, most recent epidemiological studies use sophisticated time series and distributed non-linear lag models to address this (Gasparrini, 2021; Gasparrini and Armstrong, 2010). Future research should use these methods where possible. Fourth, this study did not explore the effect of minimum temperatures, which may be relevant in the association between heat and mental health, particularly its importance on the impact of sleep (Bundo et al., 2023; Chevance et al., 2024; Jin et al., 2023; Wang et al., 2023). Lastly, given the several theoretical bio-psycho-social causal pathways between temperature exposure and mental health (Berry et al., 2010), future research should include individual biological, psychological and social factors (e.g., sex, education and employment), and symptom-related factors (e.g., prescription medication and sleep) as covariates or mediators to better understand underlying mechanisms. We have demonstrated that EMA is an exciting approach for exploring these individual-level factors in large clinical cohorts.

## Conclusions

Given rising temperatures in the context of global climate change, understanding how people with mood disorders are affected by heat is important. Our study provides evidence of a link between mean and maximum temperature and depressive and manic symptoms, suggesting people with bipolar disorder are at higher risk of manic episodes during hotter temperatures whilst people with depression and bipolar disorder experience a reduction in depressive symptoms. However, the relationship between heat and mood disorders is complex and future work needs to understand who is more at risk and the underlying mechanistic pathways.

We demonstrate the value of using novel data and EMA methods to inform our understanding of the link between climate and mental health, but improved data collection is needed to realise their full potential. Future work could use novel EMA data to explore individual-level temperature exposures alongside climate and socioeconomic covariates and interacting effects of individual-level activity and symptom-relevant factors, particularly prescription medication and sleep. However, significant limitations to EMA data including attrition and selection bias should be addressed.

## Data availability statement

Please contact authors for data. An anonymised dataset is available from juli Health, given appropriate permissions.

## Authors statement

PC, JFH, AK, DPJO, RT, ELL and JD were involved in the conception and design of the work.

AK and JFH were involved in the acquisition of data. PC analysed data.

All authors were involved in the interpretation of data. PC drafted the manuscript.

All authors contributed to drafts of the manuscript.

All authors approved the final draft of the version to be published.

