## Supplementary Material for "The association between outdoor ambient temperature and depression and mania: an ecological momentary assessment study"

### Supplement A

**Figure A1:** QQ plots for PHQ-8 and ASRM multi-level linear models

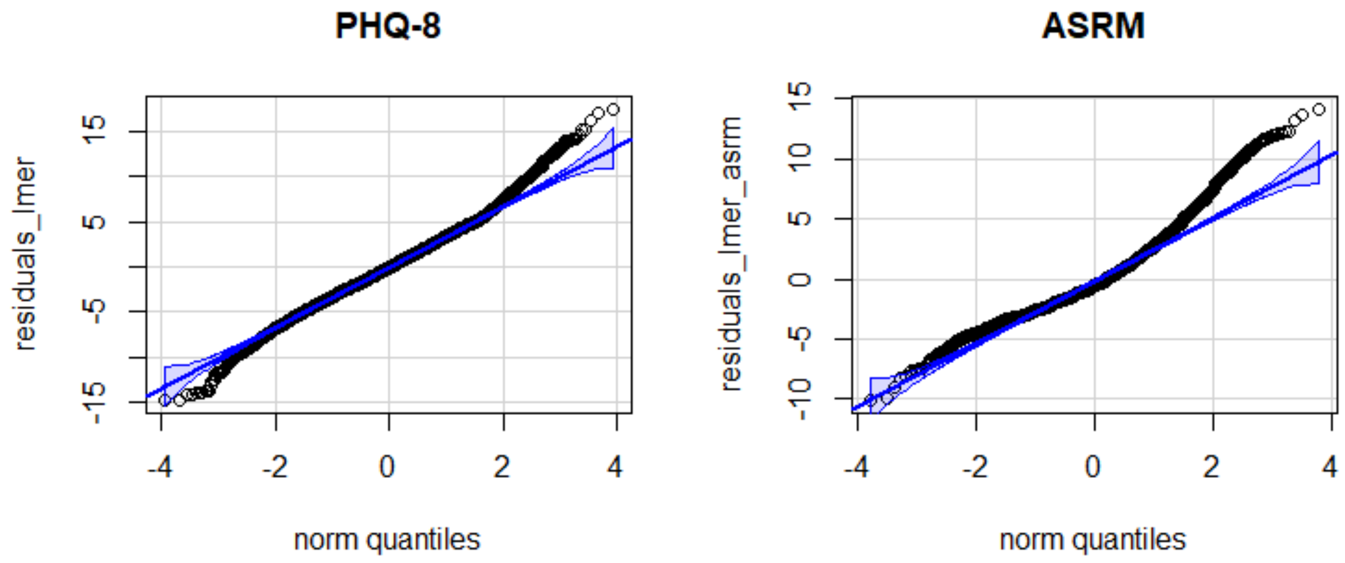


**Table A1**: Model fit comparison using AIC and BIC

| | AIC | BIC |
|---|---|---|
| **PHQ** | | |
| **Poisson (mixed effects)** | 80431.2 | 80453.6 |
| **Negative binomial (mixed effects)** | 80376.3 | 80406.1 |
| **ASRM** | | |
| **Poisson (mixed effects)** | 34549.1 | 34569.4 |
| **Negative binomial (mixed effects)** | 32409.8 | 32436.9 |

### Supplement B

**Table B1:** Geographical distribution of the total cohort of participants

| **Region** | **Percentage of total cohort (%)** |
|---|---|
| North America | 83.8 |
| Europe | 8.7 |
| Central America | 0.8 |
| Asia | 0.7 |
| Australia and New Zealand | 0.4 |
| South America | 0.3 |
| Africa | 0.2 |
| The Middle East | 0.2 |
| Missing | 5.0 |
| **Hemisphere** | |
| North | 94.3 |
| South | 0.7 |
| Missing | 5.0 |

### Supplement C

**Table C1**: Attrition table for participants in the depressive symptoms cohort who had complete-case data for all exposure, outcome, and confounding variables (listed below)

| | **Complete-case*** | **Missing** | **OR** | **95% CI** | **p** |
|---|---|---|---|---|---|
| **N_id** | 4000 | 1107 | | | |
| **Gender** | | | | | |
| Female | 8135 (78.0) | 1850 (77.1) | Ref | Ref | Ref |
| Male | 1535 (14.7) | 381 (15.9) | 1.09 | 0.96 to 1.23 | 0.163 |
| Non-binary or Transgender | 753 (7.2) | 167 (7.0) | 0.98 | 0.82 to 1.16 | 0.779 |
| Missing | 0 (0.0) | 3 (0.1) | - | - | - |
| **Age** | | | | | |
| <45 years | 5549 (53.2) | 1733 (72.2) | Ref | Ref | Ref |
| 45+ years | 1370 (13.1) | 368 (15.3) | 0.86 | 0.76 to 0.98 | **0.020** |
| Missing | 3504 (33.6) | 300 (12.5) | - | - | - |
| **Ethnicity** | | | | | |
| American Indian, Alaska Native, Native Hawaiian or Other Pacific Islander | 34 (0.3) | 1 (0.0) | Ref | Ref | Ref |
| Asian | 100 (1.0) | 11 (0.5) | 3.74 | 0.69 to 69.6 | 0.214 |
| Black-or-African American | 248 (2.4) | 20 (0.8) | 2.74 | 0.54 to 50.0 | 0.332 |
| White | 2200 (21.1) | 174 (7.2) | 2.69 | 0.58 to 47.9 | 0.330 |
| Missing | 7841 (75.2) | 2195 (91.4) | - | - | - |
| **Hispanic** | | | | | |
| No | 2358 (22.6) | 182 (7.6) | Ref | Ref | Ref |
| Yes | 332 (3.2) | 27 (1.1) | 1.05 | 0.68 to 1.58 | 0.807 |
| Missing | 7733 (74.2) | 2192 (91.3) | - | - | - |
| **Diagnosis = Bipolar Disorder** | 4707 (45.2) | 1109 (46.2) | 1.04 | 0.95 to 1.14 | 0.361 |
| **Comorbid = Yes** | 1384 (13.3) | 126 (5.2) | 0.36 | 0.30 to 0.44 | **<0.001** |
| **Duration of diagnosis** | | | | | |
| < 1 year | 882 (8.5) | 255 (10.6) | Ref | Ref | Ref |
| 1-5 years | 3085 (29.6) | 738 (30.7) | 0.83 | 0.71 to 0.97 | **0.021** |
| > 5 years | 6456 (61.9) | 1408 (58.6) | 0.75 | 0.65 to 0.88 | **<0.001** |
| **Temp 14-day mean °C (mean (SD))** | 14.11 (9.51) | 17.36 (10.17) | 1.04 | 1.03 to 1.04 | **<0.001** |
| **Temp 14-day max °C (mean (SD))** | 23.21 (9.95) | 22.37 (10.58) | 0.99 | 0.99 to 1.00 | **<0.001** |
| **Mean All-year-round Temp °C (mean (SD))** | 14.28 (7.66) | 15.95 (8.55) | 1.03 | 1.02 to 1.03 | **<0.001** |
| **Season (n(%))** | | | | | |
| Autumn | 2519 (24.2) | 781 (32.5) | Ref | Ref | Ref |
| Spring | 2202 (21.1) | 312 (13.0) | 0.46 | 0.40 to 0.53 | **<0.001** |
| Summer | 2366 (22.7) | 852 (35.5) | 1.16 | 1.04 to 1.30 | **0.009** |
| Winter | 3336 (32.0) | 456 (19.0) | 0.44 | 0.39 to 0.50 | **<0.001** |
| **Mean Season Temp °C (mean (SD))** | 14.02 (8.92) | 16.51 (9.54) | 1.03 | 1.03 to 1.04 | **<0.001** |
| **PHQ-8 total score (mean (SD))** | 12.18 (6.23) | 13.51 (6.12) | 1.04 | 1.03 to 1.04 | **<0.001** |

| | | | | | |
|---|---|---|---|---|---|
| **PHQ-8s Completed (mean (SD))** | 7.71 (8.19) | 4.43 (5.97) | 0.92 | 0.92 to 0.93 | **<0.001** |
| **Humidity 14-day (%) (mean (SD))** | 66.18 (13.64) | 69.73 (15.56) | 1.02 | 1.02 to 1.02 | **<0.001** |
| **Sunshine counts 14-day (mean (SD))** | 6.64 (5.45) | 8.91 (7.57) | 1.06 | 1.04 to 1.07 | **<0.001** |
| **Air Quality 14-day (mean (SD))** | 44.49 (21.55) | 41.03 (19.75) | 0.99 | 0.99 to 0.99 | **<0.001** |
| **Step count 14-day (mean(SD))** | | | | | |
| <1000 | 914 (8.8) | 361 (15.0) | Ref | Ref | Ref |
| 1000-3000 | 3262 (31.3) | 650 (27.1) | 0.51 | 0.44 to 0.59 | **<0.001** |
| 3001-5000 | 2616 (25.1) | 438 (18.2) | 0.42 | 0.36 to 0.50 | **<0.001** |
| 5001-7000 | 1512 (14.5) | 264 (11.0) | 0.44 | 0.37 to 0.53 | **<0.001** |
| 7001-9000 | 800 (7.7) | 144 (6.0) | 0.46 | 0.37 to 0.56 | **<0.001** |
| >9000 | 585 (5.6) | 123 (5.1) | 0.532 | 0.42 to 0.67 | **<0.001** |
| NA | 568 (5.4) | 351 (14.6) | - | - | - |

* Complete case is defined as complete for the following variables: 14-day average temperature, 14-day maximum temperature, PHQ-8 score, gender, 14-day humidity, 14-day sunshine count, 14-day air quality.
Significant associations are indicated with **bolded p values**

**Table C2**: Attrition table for the participants in the manic symptoms cohort who had complete-case data for all exposure, outcome, and confounding variables (listed below)

| | Complete-case* | missing | OR | 95% CI | p |
|---|---|---|---|---|---|
| **N_id** | 2132 | 554 | | | |
| **Gender** | | | | | |
| Female | 4160 (79.9) | 941 (76.1) | Ref | Ref | Ref |
| Male | 680 (13.1) | 198 (16.0) | 1.29 | 1.08 to 1.53 | **0.004** |
| Non-binary or Transgender | 364 (7.0) | 97 (7.8) | 1.18 | 0.93 to 1.48 | 0.171 |
| Missing | 0 (0.0) | 1 (0.1) | - | - | - |
| **Age** | | | | | |
| <45 years | 3175 (61.0) | 978 (79.1) | Ref | Ref | Ref |
| 45+ years | 506 (9.7) | 121 (9.8) | 0.78 | 0.63 to 0.96 | **0.019** |
| Missing | 1523 (29.3) | 138 (11.2) | - | - | - |
| **Ethnicity** | | | | | |
| American Indian, Alaska Native, Native Hawaiian or Other Pacific Islander | 29 (0.6) | 1 (0.1) | Ref | Ref | Ref |
| Asian | 41 (0.8) | 5 (0.4) | 3.54 | 0.53 to 69.7 | 0.260 |
| Black-or-African American | 166 (3.2) | 10 (0.8) | 1.75 | 0.32 to 32.7 | 0.601 |
| White | 1106 (21.3) | 92 (7.4) | 2.41 | 0.51 to 43.2 | 0.389 |
| Missing | 3862 (74.2) | 1129 (91.3) | - | - | - |
| **Hispanic** | | | | | |
| No | 1196 (23.0) | 98 (7.9) | Ref | Ref | Ref |
| Yes | 200 (3.8) | 12 (1.0) | 0.73 | 0.38 to 1.31 | 0.323 |
| Missing | 3808 (73.2) | 1127 (91.1) | - | - | - |
| **Diagnosis = Bipolar Disorder** | 5204 (100.0) | 1237 (100.0) | - | - | - |
| **Comorbid = Yes** | 399 (7.7) | 38 (3.1) | 0.38 | 0.27 to 0.53 | <**0.001** |
| **Duration of diagnosis** | | | | | |
| < 1 year | 612 (11.8) | 171 (13.8) | Ref | Ref | Ref |
| 1-5 years | 1588 (30.5) | 372 (30.1) | 0.84 | 0.69 to 1.03 | 0.090 |
| > 5 years | 3004 (57.7) | 694 (56.1) | 0.83 | 0.69 to 1.00 | 0.050 |
| **Temp 14-day mean °C (mean (SD))** | 15.37 (9.48) | 18.23 (10.19) | 1.03 | 1.03 to 1.04 | **<0.001** |
| **Temp 14-day max °C (mean (SD))** | 24.30 (9.85) | 23.43 (10.59) | 0.99 | 0.99 to 1.00 | **0.006** |
| **Mean All-year-round Temp °C (mean (SD))** | 15.53 (7.50) | 16.73 (8.53) | 1.02 | 1.01 to 1.03 | **<0.001** |
| **Season (n(%))** | | | | | |
| Autumn | 1165 (22.4) | 339 (27.4) | Ref | Ref | Ref |
| Spring | 1203 (23.1) | 195 (15.8) | 0.56 | 0.46 to 0.68 | **<0.001** |
| Summer | 1457 (28.0) | 486 (39.3) | 1.15 | 0.98 to 1.34 | 0.092 |
| Winter | 1379 (26.5) | 217 (17.5) | 0.541 | 0.45 to 0.65 | **<0.001** |
| **Mean Season Temp °C (mean (SD))** | 15.29 (8.92) | 17.43 (9.68) | 1.03 | 1.02 to 1.03 | **<0.001** |
| **ASRM total score (mean (SD))** | 4.29 (3.90) | 4.53 (3.85) | 1.02 | 1.00 to 1.03 | 0.054 |
| **ASRMs Completed (mean (SD))** | 7.68 (8.33) | 4.55 (6.08) | 0.93 | 0.92 to 0.94 | **<0.001** |
| **Humidity 14-day (%) (mean (SD))** | 64.21 (14.22) | 68.87 (16.09) | 1.02 | 1.02 to 1.03 | **<0.001** |
| **Sunshine counts 14-day (mean (SD))** | 6.73 (5.89) | 9.71 (7.61) | 1.06 | 1.04 to 1.08 | **<0.001** |

| | | | | | |
|---|---|---|---|---|---|
| **Air Quality 14-day (mean (SD))** | 41.67 (17.31) | 40.31 (17.19) | 1.00 | 0.99 to 1.00 | **0.021** |
| **Step count 14-day (mean(SD))** | | | | | |
| <1000 | 442 (8.5) | 153 (12.4) | Ref | Ref | Ref |
| 1000-3000 | 1705 (32.8) | 349 (28.2) | 0.59 | 0.48 to 0.74 | **<0.001** |
| 3001-5000 | 1271 (24.4) | 234 (18.9) | 0.53 | 0.42 to 0.67 | **<0.001** |
| 5001-7000 | 760 (14.6) | 134 (10.8) | 0.51 | 0.39 to 0.66 | **<0.001** |
| 7001-9000 | 371 (7.1) | 81 (6.5) | 0.63 | 0.47 to 0.85 | **0.003** |
| >9000 | 289 (5.6) | 65 (5.3) | 0.65 | 0.47 to 0.90 | **0.010** |
| NA | 366 (7.0) | 221 (17.9) | | | |

* Complete case is defined as complete for the following variables: 14-day average temperature, 14-day maximum temperature, PHQ-8 score, gender, 14-day humidity, 14-day sunshine count, 14-day air quality.
Significant associations are indicated with **bolded p values**

**Supplement D**

**Table D** (below) presents descriptive results for *all observations* in the depressive and manic symptoms cohort. We present the following across the four seasons: numbers of PHQ-8 and ASRM questionnaires completed, PHQ-8 and ASRM symptom scores, numbers of temperature recordings, and temperature values

| | Spring | Summer | Autumn | Winter | Total |
|---|---|---|---|---|---|
| **Depressive Symptoms Cohort** | | | | | |
| N | 1,204 | 1,193 | 1,279 | 1,800 | 4,000 |
| **PHQ-8 completion** | | | | | |
| Total scores completed | 2,202 | 2,366 | 2,519 | 3,336 | 10,423 |
| Min-max | 1-7 | 1-7 | 1-11 | 1-12 | 1-37 |
| Median | 1 | 1 | 1 | 1 | 1 |
| Mean (SD) | 1.61 (1.05) | 1.68 (1.19) | 1.76 (1.51) | 1.62 (1.21) | 2.61 (3.44) |
| **PHQ-8 scores** | | | | | |
| Min-max | 0-24 | 0-24 | 0-24 | 0-24 | 0-24 |
| Median | 12 | 11 | 12 | 12 | 12 |
| Mean (SD) | 11.90 (6.21) | 11.77 (6.11) | 12.22 (6.28) | 12.62 (6.24) | 12.18 (6.23) |
| **Daily temp recordings** | | | | | |
| Total recordings | 24,185 | 24,618 | 31,133 | 33,309 | 113,245 |
| Min-max | 1-109 | 1-145 | 0-182 | 1-180 | 1-616 |
| Median | 25 | 30 | 31 | 24 | 11 |
| Mean (SD) | 35.18 (29.75) | 38.62 (31.03) | 40.33 (37.84) | 36.47 (36.44) | 28.31 (50.45) |
| **Daily Temp $^{o}$C** | | | | | |
| Mean (SD) | 13.70 (7.03) | 20.65 (5.71) | 16.32 (6.18) | 8.42 (7.40) | 14.22 (8.12) |
| Mean min (SD) | 10.67 (7.69) | 19.33 (4.40) | 13.51 (6.52) | 3.43 (7.18) | 11.01 (8.90) |
| Absolute min | -29 | 6 | -16 | -35 | -35 |
| Mean max (SD) | 16.23 (7.66) | 29.65 (4.67) | 21.19 (7.54) | 8.46 (7.46) | 17.99 (10.58) |
| Absolute max | 41 | 46 | 44 | 31 | 46 |
| **Manic Symptoms Cohort** | | | | | |
| N | 689 | 815 | 610 | 790 | 2132 |
| **ASRM completion** | | | | | |
| Total scores completed | 1,203 | 1,457 | 1,165 | 1,379 | 5,204 |
| Min-max | 1-7 | 1-7 | 1-11 | 1-10 | 1-37 |
| Median | 1 | 1 | 1 | 1 | 1 |
| Mean (SD) | 1.48 (0.95) | 1.52 (1.04) | 1.69 (1.47) | 1.44 (1.06) | 2.44 (3.36) |
| **ASRM scores** | | | | | |
| Min-max | 0-20 | 0-20 | 0-20 | 0-20 | 0-20 |
| Median | 4 | 4 | 3 | 3 | 3 |
| Mean (SD) | 4.51 (3.92) | 4.45 (3.97) | 4.03 (3.76) | 4.15 (3.94) | 4.29 (3.90) |
| **Daily temp recordings** | | | | | |
| Total recordings | 1,203 | 12,981 | 14,793 | 13,288 | 53,366 |
| Min-max | 1-98 | 1-92 | 1-182 | 1-137 | 1-614 |
| Median | 6 | 6 | 6 | 6 | 6 |
| Mean (SD) | 13.76 (15.49) | 13.34 (16.04) | 15.33 (23.53) | 12.48 (15.88) | 25.03 (49.15) |
| **Daily Temp $^{o}$C** | | | | | |
| Mean (SD) | 14.21 (6.83) | 21.24 (5.99) | 16.50 (6.16) | 9.72 (7.56) | 15.50 (7.95) |
| Mean Min (SD) | 11.41 (7.71) | 19.56 (4.51) | 13.66 (6.42) | 4.15 (7.44) | 12.27 (8.73) |
| Absolute min | -16 | 6 | -15 | -34 | -34 |
| Mean Max (SD) | 16.35 (7.60) | 30.02 (4.69) | 21.35 (7.64) | 8.51 (7.85) | 19.22 (10.71) |
| Absolute max | 40 | 45 | 43 | 31 | 45 |

### Supplement E

**Table E1:** Sensitivity analyses for temperature (mean, maximum) and ***depressive*** symptoms scores (PHQ-8)

| | **Model S1**[1] | | | **Model S2**[2] | | | **Model S3**[3] | | |
|---|---|---|---|---|---|---|---|---|---|
| | **N** | **IRR** | **95% CI** | **N** | **IRR** | **95% CI** | **N** | **IRR** | **95% CI** |
| **Mean temperature** | | | | | | | | | |
| **Overall** | | | | | | | | | |
| All seasons | 4000 | 0.998 | 0.997 - 1.000* | 2486 | 0.997 | 0.996 - 0.998*** | 3710 | 0.998 | 0.997 - 0.999*** |
| **Season** | | | | | | | | | |
| Spring | 1204 | 0.991 | 0.986 - 0.996*** | 1041 | 0.995 | 0.991 - 0.998** | 1132 | 0.996 | 0.993 - 1.000* |
| Summer | 1193 | 0.991 | 0.983 - 0.999* | 465 | 0.992 | 0.983 - 1.000 | 1113 | 0.994 | 0.998 - 1.000 |
| Autumn | 1278 | 1.001 | 0.998 - 1.005 | 867 | 0.999 | 0.995 - 1.002 | 1143 | 0.999 | 0.996 - 1.002 |
| Winter | 1800 | 1.005 | 1.000 - 1.009* | 1137 | 1.001 | 0.998 - 1.004 | 1693 | 1.002 | 0.999 - 1.004 |
| **Maximum temperature** | | | | | | | | | |
| **Overall** | | | | | | | | | |
| All seasons | 4000 | 0.995 | 0.994 - 0.997*** | 2486 | 0.995 | 0.994 - 0.996*** | 3710 | 0.996 | 0.995 - 0.997*** |
| **Season** | | | | | | | | | |
| Spring | 1204 | 0.989 | 0.985 - 0.993*** | 1041 | 0.992 | 0.989 - 0.996*** | 1132 | 0.994 | 0.990 - 0.997*** |
| Summer | 1193 | 0.989 | 0.983 - 0.995*** | 465 | 0.993 | 0.986 - 1.001 | 1113 | 0.990 | 0.985 - 0.996** |
| Autumn | 1278 | 0.996 | 0.993 - 0.999* | 867 | 0.994 | 0.991 - 0.998** | 1143 | 0.996 | 0.993 - 0.999** |
| Winter | 1800 | 0.995 | 0.991 - 0.998** | 1137 | 0.998 | 0.995 - 1.001 | 1693 | 0.997 | 0.995 - 1.000* |

[1] adjusted for gender, humidity, sunshine, air pollution, individual temperature norms
[2] adjusted for gender, humidity, sunshine, air pollution, age
[3] adjusted for gender, humidity, sunshine, air pollution, step count
N = number of participants
P values *<0.05**<0.01 ***<0.001

**Table E2:** Sensitivity analyses for temperature (mean, maximum) and ***manic*** symptoms scores (ASRM)

| | Model S1[1] | | | Model S2[2] | | | Model S3[3] | | |
|---|---|---|---|---|---|---|---|---|---|
| | **N** | **IRR** | **95% CI** | **N** | **IRR** | **95% CI** | **N** | **IRR** | **95% CI** |
| **Mean temperature** | | | | | | | | | |
| **Overall** | | | | | | | | | |
| All seasons | 2132 | 1.007 | 1.002 - 1.011** | 1389 | 1.002 | 0.998 - 1.005 | 1958 | 1.004 | 1.001 - 1.008** |
| **Season** | | | | | | | | | |
| Spring | 689 | 0.996 | 0.982 - 1.010 | 660 | 1.001 | 0.993 - 1.009 | 636 | 1.004 | 0.996 - 1.013 |
| Summer | 815 | 1.004 | 0.979 - 1.030 | 236 | 1.012 | 0.989 - 1.035 | 753 | 1.003 | 0.989 - 1.017 |
| Autumn | 609 | 1.019 | 1.006 - 1.032** | 423 | 1.006 | 0.996 - 1.016 | 539 | 1.010 | 1.001 - 1.020* |
| Winter | 790 | 1.011 | 0.995 - 1.028 | 647 | 1.003 | 0.994 - 1.011 | 746 | 1.004 | 0.995 - 1.012 |
| **Maximum temperature** | | | | | | | | | |
| **Overall** | | | | | | | | | |
| All seasons | 2132 | 1.005 | 1.001 - 1.009* | 1389 | 1.001 | 0.998 - 1.005 | 1958 | 1.003 | 1.000 - 1.006 |
| **Season** | | | | | | | | | |
| Spring | 689 | 1.000 | 0.989 - 1.011 | 660 | 1.002 | 0.994 - 1.010 | 636 | 1.004 | 0.996 - 1.012 |
| Summer | 815 | 0.997 | 0.980 - 1.015 | 236 | 0.999 | 0.977 - 1.022 | 753 | 0.995 | 0.980 - 1.010 |
| Autumn | 609 | 1.010 | 0.998 - 1.021 | 423 | 1.007 | 0.997 - 1.016 | 539 | 1.007 | 0.998 - 1.017 |
| Winter | 790 | 1.002 | 0.990 - 1.014 | 647 | 1.001 | 0.993 - 1.009 | 746 | 1.000 | 0.992 - 1.008 |

[1] adjusted for gender, humidity, sunshine, air pollution, individual temperature norms
[2] adjusted for gender, humidity, sunshine, air pollution, age
[3] adjusted for gender, humidity, sunshine, air pollution, step count
N_id = number of participants
N_obs = number of observations (i.e. PHQ-8 or ASRM symptoms score questionnaires completed)
P values *<0.05**<0.01 ***<0.001

**Table E3:** Results of interaction analysis for gender and age

| | Depressive symptoms cohort | | Manic symptoms cohort | |
|---|---|---|---|---|
| | **Interaction estimate** | **95%CI** | **Interaction estimate** | **95%CI** |
| **Mean temperature** | | | | |
| **Gender** | | | | |
| Female | *Ref* | *Ref* | *Ref* | *Ref* |
| Male | 0.999 | 0.996 - 1.002 | 0.997 | 0.988 - 1.006 |
| Non-binary or transgender | 1.001 | 0.997 - 1.005 | 0.999 | 0.987-1.011 |
| **Age** | | | | |
| <45 | *Ref* | *Ref* | *Ref* | *Ref* |
| ≥45 | 0.999 | 0.995-1.002 | 0.999 | 0.988-1.009 |
| **Maximum temperature** | | | | |
| **Gender** | | | | |
| Female | *Ref* | *Ref* | *Ref* | *Ref* |
| Male | 1.000 | 0.997-1.003 | 0.998 | 0.990-1.007 |
| Non-binary or transgender | 1.002 | 0.999-1.006 | 0.995 | 0.984-1.006 |
| **Age** | | | | |
| <45 | *Ref* | *Ref* | *Ref* | *Ref* |
| ≥45 | 0.999 | 0.996-1.002 | 0.998 | 0.988-1.008 |

**Table E4:** Results of the depressive symptom cohort interaction analysis by diagnosis

| | Mean temperature | | Maximum temperature | |
|---|---|---|---|---|
| | **Interaction estimate** | **95%CI** | **Interaction estimate** | **95%CI** |
| **Diagnosis** | | | | |
| Depression | *Ref* | *Ref* | *Ref* | *Ref* |
| Bipolar disorder | 0.999 | 0.996-1.001 | 0.998 | 0.996-1.000 |
